# AnnotX: An Edge-powered Laparoscopic Video Annotation Platform

**DOI:** 10.64898/2026.05.11.26352930

**Authors:** Marzie Lafouti, Liane S. Feldman, Amir Hooshiar

## Abstract

Accurate and objective evaluation of surgical skill and performance is critical for advancing training and improving patient outcomes. Current assessment methods increasingly rely on video analytics and depend on labor-intensive, frame-by-frame manual annotation by experts. In this work we developed a surgical video annotation platform (AnnotX) that used a Python backend running a pretrained promptable video segmentation foundation model, i.e., Segment Anything 3 (SAM 3) for per frame segmentation and temporal segment propagation. With a few interactions per class, the model generated a high-quality mask on a key frame and propagated it through the sequence. The platform automatically exported per-class binary masks and color overlays for every frame, together with deterministic metadata and a standardized study folder structure to support auditability and downstream analysis. On deidentified laparoscopic surgery videos, the system processed typical clips in minutes and reduced expert annotation time from hours to minutes without task-specific fine-tuning. We also benchmarked multiple SAM variants (SAM 2, MedSAM 2, and SAM 3) on the CholecSeg8K dataset, and showed AnnotX with a SAM 3 backbone outperformed alternatives. It exhibited a mean IoU of 0.884 and mean Dice of 0.924 across 101 annotated sequences. By being free, practical, and lightweight to deploy, AnnotX aims to accelerate reproducible surgical dataset creation and provides a step toward scalable, video-based performance evaluation in training and quality-improvement settings.

## 1 Introduction

### 1.1 Background and clinical need

Objective, scalable assessment of surgical skill increasingly relies on the analysis of operative video, however, current workflows remain constrained by labor-intensive, frame-by-frame annotation performed by experts. This burden limits dataset size, slows the cycle of feedback to trainees, and introduces inter-rater variability that complicates reproducibility and benchmarking. The field of Surgical Data Science has repeatedly called for standardized, data-driven pipelines that reduce annotator effort while preserving clinical relevance and auditability [16]. Recent clinical work from the same ecosystem shows that structured intraoperative video self-review is feasible and supported by valid assessment tools, underscoring the need for efficient capture, review, and annotation workflows [3, 4]. In the laparoscopic setting, complex visual conditions including occlusion, smoke, instrument specularities, blood, and rapid viewpoint change further increase the cost of pixel-accurate labeling and hinder consistent large-scale curation [20]. As a result, despite the ubiquity of recorded procedures, many groups remain bottlenecked by annotation rather than modeling, and skill evaluation pipelines remain difficult to scale beyond small cohorts. Complementary approaches aim to reduce review burden by producing saliency-guided surgical highlight reels that expose the most influential frames and time points for automated skill inference [12]. Pixel-accurate labels clearly enable clinically meaningful guidance: Madani et al. delineated “Go/No-Go” zones in the hepatocystic triangle via semantic segmentation during laparoscopic cholecystectomy: an approach that depends on reliable mask creation [15].

### 1.2 State-of-the-art (SOTA)

A large body of prior work has explored surgical scene understanding and skill assessment from video, including phase recognition and tool presence (e.g., EndoNet/Cholec80) [19], and benchmarking for instrument segmentation and tracking (e.g., EndoVis 2017) [2]. Related work has applied Siamese-based instrument tracking in ophthalmic surgery, an approach that can be generalized to multilabel laparoscopic scenes [10]. These efforts established common tasks, public datasets, and evaluation metrics that significantly advanced reproducibility in surgical data science. Motivated by this emphasis on reproducibility, we quantitatively benchmark AnnotX using SAM 3 on the public CholecSeg8K semantic segmentation dataset [7] using standard overlap metrics (IoU and Dice), in addition to demonstrating qualitative results on de-identified clinical videos. Collectively, education studies leveraging recorded video, AI guidance reliant on precise masks, and robotics-driven workflows motivate a surgeon-oriented platform that produces temporally consistent, auditable annotations with minimal expert effort [3, 15, 18]. However, most of these studies relied on limited datasets often <100 procedures and required exhaustive frame-by-frame manual annotation to achieve sufficient accuracy. As a result, scalability remained a major limitation: dataset expansion was constrained by the enormous labeling cost and inter-observer variability inherent to manual pixel-level annotation in complex laparoscopic scenes. Moreover, many benchmarks focused on isolated subtasks such as tool presence, phase classification, or binary segmentation rather than complete, multi-class surgical environments where tools and tissues interact dynamically.

The recent emergence of foundation models for segmentation has dramatically changed the annotation landscape. The original Segment Anything Model (SAM) introduced by Meta demonstrated a general-purpose image segmentation framework capable of producing object masks from simple prompts such as bounding boxes, points, or text [9]. SAM’s success stems from large-scale pretraining and strong zero-shot generalization, but the base model was designed for static 2D images and does not directly enforce temporal consistency across video frames. To adapt this paradigm for medical imaging, MedSAM fine-tunes SAM on diverse medical data (e.g., CT, MRI, ultrasound, and endoscopy), improving delineation of clinical structures and reducing prompt sensitivity [14]. Extending promptable segmentation to video, SAM 2 introduced a streaming-memory mechanism that propagates masks temporally while supporting interactive correction, making it well suited to annotation scenarios where temporal coherence is essential [17]. Building on this direction, MedSAM2 variants adapt SAM 2 for universal 2D/3D medical segmentation by formulating segmentation as tracking across slices/frames and introducing memory-selection strategies (e.g., self-sorting memory) to improve robustness and cross-slice/temporal consistency [21, 14]. More recently, SAM 3 further advances the Segment Anything line as a unified model for detection, segmentation, and tracking in images and videos, improving robustness and usability across diverse settings [5]. In parallel, Med-SAM3 adapts the SAM 3 architecture for medical image and video segmentation by incorporating medical concepts and open-vocabulary text prompts to target anatomical structures more precisely [13].

While these foundation models are powerful, their deployment in surgical annotation workflows remains limited: popular open-source tools (e.g., CVAT, Label Studio) lack native promptable video propagation and are not optimized for clinician interaction [6, 1]. Together, these trends highlight an opportunity for an intuitive, surgeon-oriented platform that operationalizes foundation-model segmentation in a transparent, auditable way for real operating-room data. This surgeon-first emphasis aligns with broader translational work in surgical robotics, where usability and dependable perception are prerequisites for clinical adoption [18]. Complementary routes toward online deployment have also been explored, including accelerated mask propagation and detection-assisted prompting for laparoscopic video, as well as online interfaces that couple detector outputs with SAM-family segmentation [11, 8]. In AnnotX, we employ SAM 3 as a practical backbone: it offers promptable control and practical temporal consistency in our offline workflows, and although we run it offline rather than real-time on our setup, its computational footprint is compatible with a standard workstation and batch annotation workflows.

### 1.3 Objective and Contributions

The objective of this study is to develop and validate **AnnotX**, a hardware-light, open-source desktop annotation platform for laparoscopic videos that integrates surgeon-friendly interactions with foundation-model segmentation (pretrained SAM 3), enabling rapid creation of audit-ready segmentation datasets with minimal expert input and without model fine-tuning.

The main contributions of this work were:

1. We introduced an open-source tool for laparoscopic video segmentation that couples a surgeon-friendly graphical interface C # with a Python backend running the pretrained SAM 3 model. The system is intentionally hardware-light, allowing a standard single-GPU workstation to annotate typical clips in minutes without fine-tuning.
2. To preserve reproducibility and traceability, we propose a robust data model and deterministic dataset structure that automatically exports per-class binary masks, color overlays, and a complete JSON manifest capturing user prompts and inference parameters.
3. We apply AnnotX to de-identified laparoscopic surgery videos to demonstrate that sparse clinician interactions on selected key frames (e.g., bounding boxes and clicks) can replace dense, frame-wise labeling, substantially reducing expert effort while producing usable segmentation outputs.

## 2 Methodology

### 2.1 Platform Overview and Design Goals

AnnotX is a desktop tool that lets a clinical annotator turn a few simple prompts into per-frame masks for laparoscopic video. It comprises a Windows C# GUI for user interaction and a Python worker running SAM 3 for segmentation (Fig. 1). The worker consumes a JSON request and writes masks, overlays, and a manifest (Figs. 1, 2). A typical session involves importing a de-identified video, defining classes (tools/tissues) with a tight box and sparse clicks on a key frame, running SAM 3 to propagate masks forward, and re-seeding when drift occurs. The worker consumes these inputs via a JSON request and exports artifacts to a fixed study folder (Fig. 2) to support auditability. Exports include per-class binary masks, RGBA color overlays for visual checks, and a manifest JSON detailing the exact prompts used (Section 2.3). SAM 3 initializes from these prompts and propagates via its memory-based tracking pipeline (Section 2.2). AnnotX runs offline on a single-GPU workstation (NVIDIA GeForce RTX 4070 Ti, 12 GB VRAM, Intel i9-class CPU, 128 GB RAM). The GUI is implemented in C#, the worker uses Python (PyTorch) and a released SAM 3 checkpoint. Communication is file-based (JSON in, PNG/JSON out), keeping integration simple in clinical IT environments. We cast annotation as promptable video segmentation. Let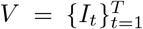 be a laparoscopic video and 𝒞 = {*c*_1_, …, *c*_*K*_} the set of user-defined semantic classes (tools/tissues). For each class *c*_*k*_, the annotator chooses one or more key frames 𝒦_*k*_ ⊂ {1, …, *T* } and supplies prompts on each key frame: a tight bounding box *b*_*k,t*_ and sparse positive/negative clicks 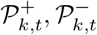.A pretrained SAM 3 model *f*_*θ*_ [5] produces a mask 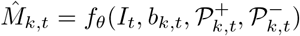 and propagates it through the sequence using its video tracking pipeline with a memory bank. The result is a per-class, per-frame binary mask set 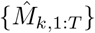.

**Fig. 1.**
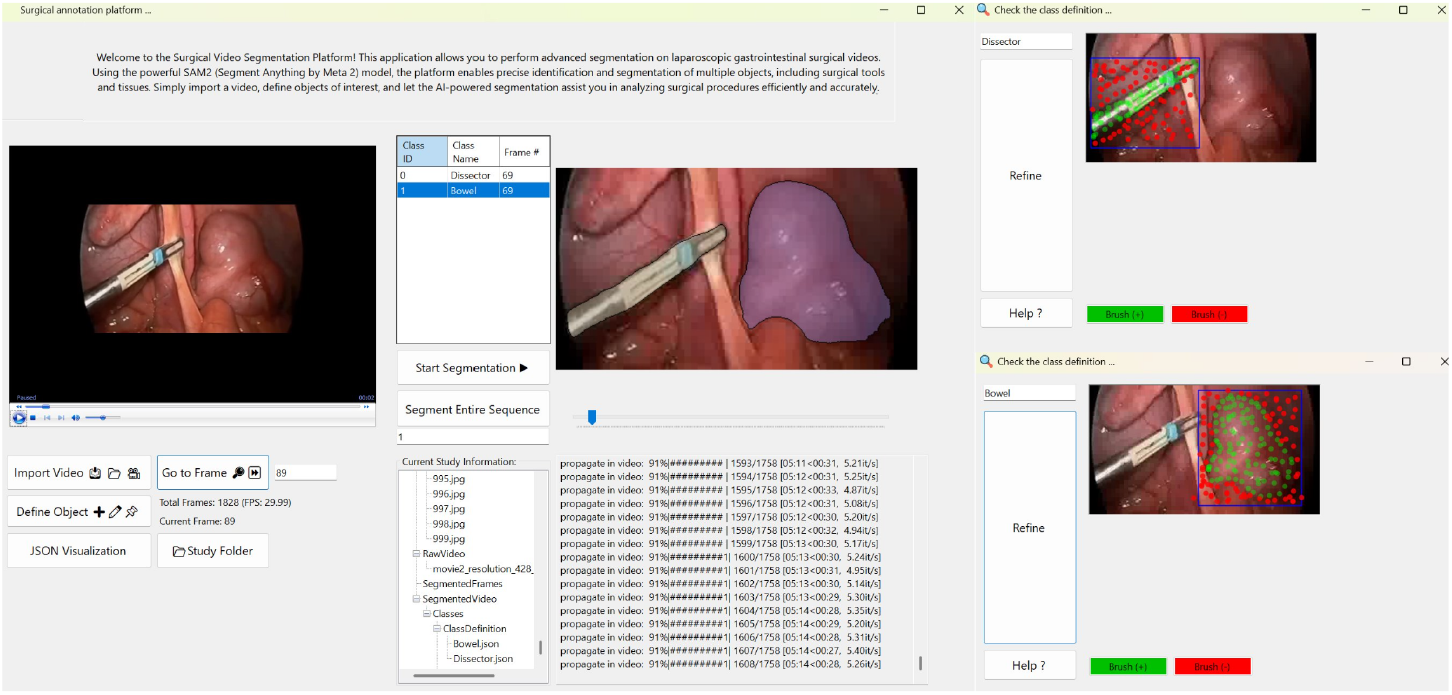
AnnotX graphical user interface (GUI). Original video view (left), live mask overlay (right), and class/prompt controls (center box) and positive (green) /negative (red) points.

**Fig. 2.**
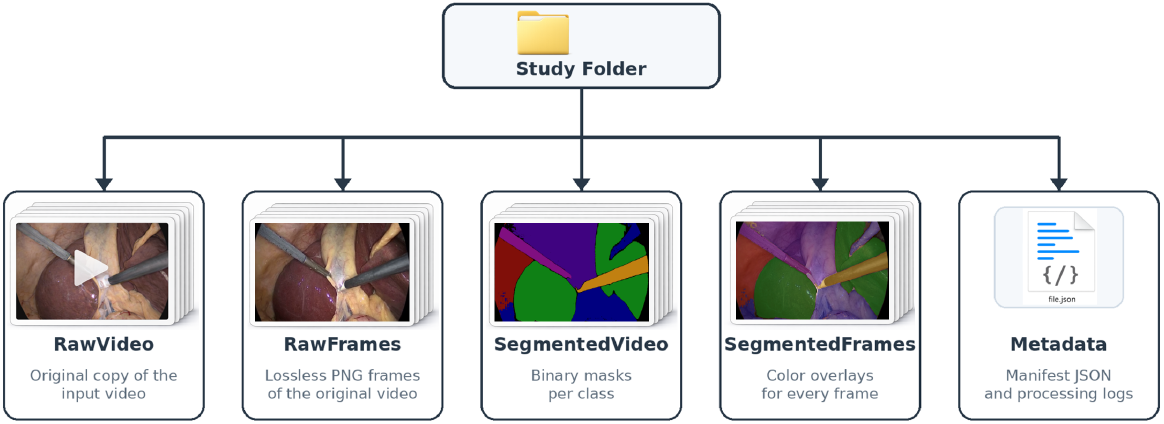
Standardized study folder hierarchy used to organize exported artifacts.

### 2.2 Segmentation and Tracking with SAM 3

Fig. 3 summarizes how SAM 3 operates in our pipeline. SAM 3 computes prompt-conditioned embeddings for mask initialization and propagates masks via a detector-tracker design with a memory bank. In our use case, the annotator provides only spatial prompts (a tight bounding box and sparse positive/negative points) on a key frame to initialize the target mask. SAM 3 then propagates masks over time using a tracker coupled to a memory bank, which helps maintain temporal consistency through occlusion, illumination change, and moderate motion. When drift is detected, the annotator re-seeds the object on the first offending frame with a small prompt update and AnnotX re-runs forward propagation from that frame. SAM 2 introduced memory-based mask propagation for video [17], and SAM 3 further advances this line with unified detection/segmentation/tracking for videos [5]. In practice, annotation follows a simple loop: seed a class on a selected key frame by the user with box and points, propagate forward, and re-seed on the first failing frame when drift is observed; AnnotX then re-runs forward and overwrites masks for that class only on frames *t*^*′*^ … *T*, leaving earlier frames unchanged (Fig. 1).

**Fig. 3.**
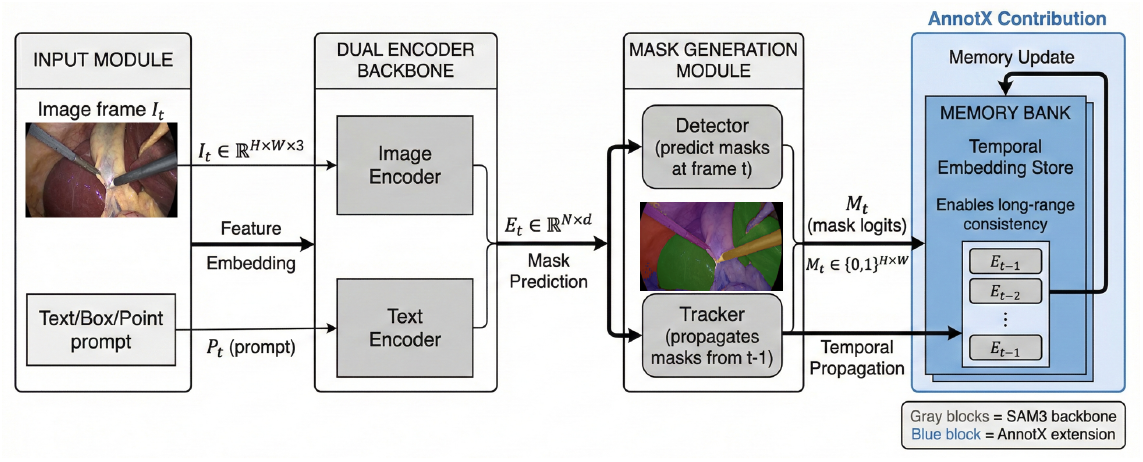
AnnotX pipeline: SAM 3-based prompted video segmentation with memory propagation.

### 2.3 Dataset Architecture and Privacy

AnnotX exports per-class binary masks, overlay visualizations, and a manifest JSON in a fixed study folder (Fig. 2). The manifest records a non-identifying video reference, class name/ID, key-frame index, segmented frame range, and the exact prompts (box and positive/negative points) used for each run (including re-seeds), enabling deterministic re-runs and auditing. AnnotX operates entirely on local, de-identified data. Frames are not transmitted outside the workstation. Manifests and logs exclude Protected Health Information and retain only non-identifying references (e.g., hashed IDs) and aggregate video properties necessary for audit and reproducibility. We focus on laparoscopic abdominal procedures recorded at a large tertiary-care teaching hospital. Videos are recorded at 30 fps and accessed securely via a clinical video management system; all data are deidentified before annotation. This work focuses on creating high-quality masks for instruments and tissues to support video-based performance evaluation and skill assessment.

## 3 Results and Discussion

### Temporal Stability

From a few prompts per class on one or more key frames, propagated masks remained temporally stable under moderate motion, contact, and lighting variation. Typical failure modes matched clinical intuition: tool exit or entry, heavy smoke, and strong specular highlights. A quick re-seed at the first offending frame restored consistency. The side-by-side overlays in the GUI (Fig. 1) made visual checks of alignment and class correctness immediate. Representative qualitative examples on CholecSeg8K are shown in Fig. 4, comparing raw frames, ground truth, and AnnotX predictions.

**Fig. 4.**
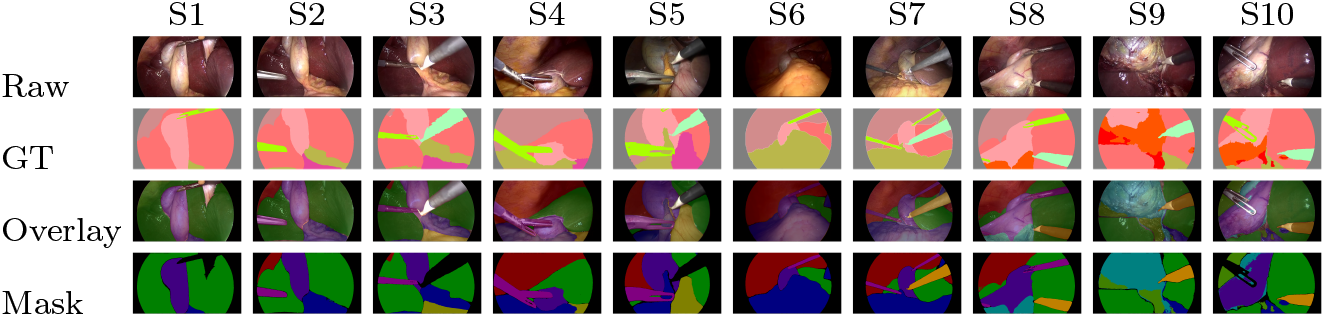
Qualitative segmentation results on CholecSeg8K for 10 samples.

### Accuracy

We assessed temporal accuracy by computing per-frame Dice between predictions and ground truth to show how accurate masks remained temporally. Fig. 5 shows mean Dice over time for each class and a combined view. Large consistently visible regions (abdominal wall, liver, fat) remain stable, whereas small/intermittent targets (cystic duct, hepatic vein, liver ligament) and fast-moving instruments (grasper, L-hook electrocautery) fluctuate under occlusion, motion blur, and specular highlights; in practice, a single re-seed typically restores performance. Therefore, additional key frames are most beneficial for rare/small targets.

**Fig. 5.**
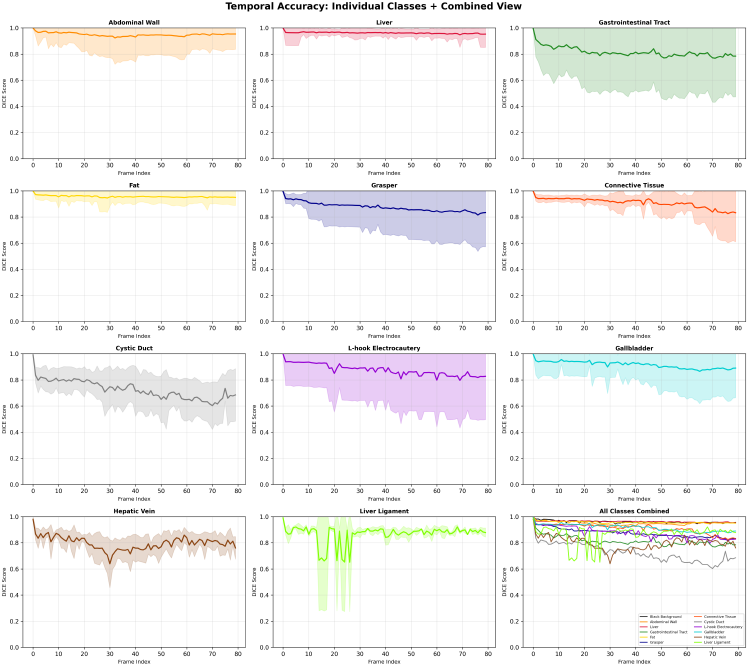
Per-class Dice over time on CholecSeg8K.

### User Effort and Workflow

The interaction loop (seed →forward propagate →re-seed if needed) kept edits minimal. Re-seeding at frame *t*^*′*^ deterministically overwrote downstream masks for that class on *t*^*′*^ … *T* while leaving earlier frames untouched, which simplified audit and avoided cascading edits. In practice, one to three key frames per class per clip sufficed for long segments without any task-specific fine-tuning. End-to-end processing including brief user interaction and exports completed in minutes for typical clips, preserving an interactive feel on a standard workstation. The fixed study folder (Fig. 2) and manifest made the outputs directly consumable by downstream pipelines.

### Benchmarking with SOTA

We benchmarked AnnotX on CholecSeg8K using fixed box-point prompting with mask-based propagation. The dataset contains 101 sequences (8,080 frames) and 11 classes excluding background. IoU and Dice are computed per frame, averaged per sequence, and reported as mean ±SD across sequences. Table 1 shows overall results: AnnotX-SAM 3 achieves the best agreement with ground truth (mIoU 0.884 ±0.067, mDice 0.924 ±0.057), slightly exceeding AnnotX-SAM 2 and outperforming AnnotX-MedSAM 2. MedSAM3-v1 is text-guided and is not directly comparable under the same box/point/mask protocol [13]. Table 2 reports per-class overlap for AnnotX-SAM 3. Large regions (liver, abdominal wall) score highest, while small/infrequent structures (cystic duct, hepatic vein, liver ligament) score lower under occlusion and ambiguous boundaries. Limitations of this work include failure cases for visually similar adjacent tissues where masks may merge or swap. In practice, adding a key frame and a few targeted positive/negative clicks resolves most cases, while domain adaptation or lightweight fine-tuning may further improve tissue differentiation. SAM 3 is not real-time on a single-GPU workstation, so we use it for offline ground-truth creation. Throughput can be improved via downscaling, mixed precision, reduced memory windows, or chunked processing.

**Table 1.** CholecSeg8K overall benchmark (mean±SD).

| Model | mIoU (↑) | mDice |
| --- | --- | --- |
| SAM 2 | 0.881±0.069 | 0.921±0.060 |
| MedSAM 2 | 0.838±0.086 | 0.891±0.075 |
| **SAM 3** | **0.884±0.067** | **0.924±0.057** |

**Table 2.**
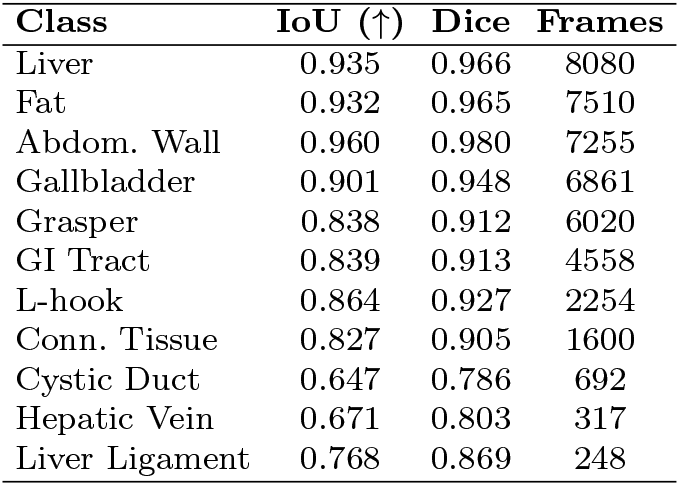
Per-class benchmarking on CholecSeg8K (AnnotX-SAM 3).

## 4 Conclusion

We presented AnnotX, an open-source surgeon-oriented desktop platform that combines a C# GUI with a SAM 3 backend to generate temporally consistent per-frame masks from sparse prompts. The system exports deterministic per-class masks, overlays, and a manifest for auditing and downstream use. On de-identified laparoscopic clips, typical cases were processed in minutes without task-specific fine-tuning, and benchmarking on CholecSeg8K achieved mIoU 0.884± 0.067 and mDice 0.924± 0.057. Future work includes adding text prompting to the GUI so that text-driven variants (e.g., MedSAM3-style workflows) can be benchmarked fairly alongside point/box/mask prompting and conducting formal usability and timing studies with surgical users.

## Data Availability

The source code for the AnnotX platform is currently being finalized. The code will be made publicly available in a dedicated repository upon peer-reviewed publication of this manuscript. In the interim, the software is available from the corresponding author upon reasonable request.

https://www.kaggle.com/datasets/newslab/cholecseg8k

## Acknowledgments

We acknowledge the Surgical Performance Enhancement and Robotics (SuPER) Centre and the McGill University Health Centre (MUHC) for institutional support. This work was supported in part by the Montreal General Hospital Foundation (MGHF) through the Mimi Dupuis Benjamin Award and the Nesbitt-McMaster Award.

## Author Contributions

M.L. conceived the study, developed the methodology and software, conducted experiments and analysis, and drafted the manuscript. L.S.F. and A.H. supervised the work and revised the manuscript.

## Ethics Statement

This research study was conducted with the formal approval of the McGill University Health Centre (MUHC) Research Ethics Board (REB), identification number: 2025-10948. Approval was obtained prior to the commencement of data collection for the institutional laparoscopic video datasets. All surgical video recordings utilized in this study were strictly de-identified to protect patient confidentiality and were handled in full compliance with established institutional privacy protocols.

## References

1. HumanSignal/label-studio (Feb 2026), https://github.com/HumanSignal/label-studio

2. Allan, M., Shvets, A., Kurmann, T., Zhang, Z., Duggal, R., Su, Y.H., Rieke, N., Laina, I., Kalavakonda, N., Bodenstedt, S., Herrera, L., Li, W., Iglovikov, V., Luo, H., Yang, J., Stoyanov, D., Maier-Hein, L., Speidel, S., Azizian, M.: 2017 Robotic Instrument Segmentation Challenge (Feb 2019). 10.48550/arXiv.1902.06426

3. Balvardi, S., Kaneva, P., Semsar-Kazerooni, K., Vassiliou, M., Al Mahroos, M., Mueller, C., Fiore, J.F., Schwartzman, K., Feldman, L.S.: Effect of video-based self-reflection on intraoperative skills: A pilot randomized controlled trial. Surgery 175(4), 1021–1028 (Apr 2024). 10.1016/j.surg.2023.11.028

4. Balvardi, S., Semsar-Kazerooni, K., Kaneva, P., Mueller, C., Vassiliou, M., Al Mahroos, M., Fiore, J.F., Schwartzman, K., Feldman, L.S.: Validity of video-based general and procedure-specific self-assessment tools for surgical trainees in laparoscopic cholecystectomy. Surgical Endoscopy 37(3), 2281–2289 (Mar 2023). 10.1007/s00464-022-09466-6

5. Carion, N., Gustafson, L., Hu, Y.T., Debnath, S., Hu, R., Suris, D., Ryali, C., Alwala, K.V., Khedr, H., Huang, A., Lei, J., Ma, T., Guo, B., Kalla, A., Marks, M., Greer, J., Wang, M., Sun, P., Rädle, R., Afouras, T., Mavroudi, E., Xu, K., Wu, T.H., Zhou, Y., Momeni, L., Hazra, R., Ding, S., Vaze, S., Porcher, F., Li, F., Li, S., Kamath, A., Cheng, H.K., Dollár, P., Ravi, N., Saenko, K., Zhang, P., Feichtenhofer, C.: SAM 3: Segment Anything with Concepts (Nov 2025). 10.48550/arXiv.2511.16719

6. Corporation, C.A.: Computer Vision Annotation Tool (CVAT). Zenodo (Apr 2023). 10.5281/zenodo.7863887

7. Hong, W.Y., Kao, C.L., Kuo, Y.H., Wang, J.R., Chang, W.L., Shih, C.S.: CholecSeg8k: A Semantic Segmentation Dataset for Laparoscopic Cholecystectomy Based on Cholec 80 (Dec 2020). 10.48550/arXiv.2012.12453

8. Hubermann, J., Lafouti, M., Levy, M., Feldman, L.S., Hooshiar, A.: Florencelapchole: Automated surgical scene inference and segmentation with natural language-guided online interface. In: Society of American Gastrointestinal and Endoscopic Surgeons (SAGES) Annual Meeting. vol. 39, p. S443 (2025). 10.1007/s00464-025-11690-9

9. Kirillov, A., Mintun, E., Ravi, N., Mao, H., Rolland, C., Gustafson, L., Xiao, T., Whitehead, S., Berg, A.C., Lo, W.Y., Dollár, P., Girshick, R.: Segment Anything (Apr 2023). 10.48550/arXiv.2304.02643

10. Lafouti, M., Ahmadi, M., Allahkaram, M., Gandomi, I., Lotfi, F., Mohammadzadeh, M., Abdi, P., Taghirad, H.D.: Surgical Instrument Tracking for Capsulorhexis Eye Surgery Based on Siamese Networks. In: 2022 10th RSI International Conference on Robotics and Mechatronics (ICRoM). pp. 196–201 (Nov 2022). 10.1109/ICRoM57054.2022.10025355

11. Lafouti, M., Feldman, L.S., Hooshiar, A.: Medsam-flow: A deep fusion methodology for robust realtime tracking of objects in laparoscopic videos. In: Proceedings of the 16th Hamlyn Symposium on Medical Robotics (HSMR) (2024). 10.31256/HSMR2024.2

12. Lafouti, M., Feldman, L.S., Hooshiar, A.: Accelerated Video-Based Surgical Skills Assessment through Saliency-Guided Surgical Highlights Reel (Nov 2025), https://openreview.net/forum?id=g9OAaDTQ27

13. Liu, A., Xue, R., Cao, X.R., Shen, Y., Lu, Y., Li, X., Chen, Q., Chen, J.: MedSAM3: Delving into Segment Anything with Medical Concepts (Nov 2025). 10.48550/arXiv.2511.19046

14. Ma, J., Yang, Z., Kim, S., Chen, B., Baharoon, M., Fallahpour, A., Asakereh, R., Lyu, H., Wang, B.: MedSAM2: Segment Anything in 3D Medical Images and Videos (Apr 2025). 10.48550/arXiv.2504.03600

15. Madani, A., Namazi, B., Altieri, M.S., Hashimoto, D.A., Rivera, A.M., Pucher, P.H., Navarrete-Welton, A., Sankaranarayanan, G., Brunt, L.M., Okrainec, A., Alseidi, A.: Artificial Intelligence for Intraoperative Guidance: Using Semantic Segmentation to Identify Surgical Anatomy During Laparoscopic Cholecystectomy. Annals of Surgery 276(2), 363 (Aug 2022). 10.1097/SLA.0000000000004594

16. Maier-Hein, L., Vedula, S.S., Speidel, S., Navab, N., Kikinis, R., Park, A., Eisenmann, M., Feussner, H., Forestier, G., Giannarou, S., Hashizume, M., Katic, D., Kenngott, H., Kranzfelder, M., Malpani, A., März, K., Neumuth, T., Padoy, N., Pugh, C., Schoch, N., Stoyanov, D., Taylor, R., Wagner, M., Hager, G.D., Jannin, P.: Surgical data science for next-generation interventions. Nature Biomedical Engineering 1(9), 691–696 (Sep 2017). 10.1038/s41551-017-0132-7

17. Ravi, N., Gabeur, V., Hu, Y.T., Hu, R., Ryali, C., Ma, T., Khedr, H., Rädle, R., Rolland, C., Gustafson, L., Mintun, E., Pan, J., Alwala, K.V., Carion, N., Wu, C.Y., Girshick, R., Dollár, P., Feichtenhofer, C.: SAM 2: Segment Anything in Images and Videos (Oct 2024). 10.48550/arXiv.2408.00714

18. Roshanfar, M., Dargahi, J., Hooshiar, A.: Design Optimization of a Hybrid-Driven Soft Surgical Robot with Biomimetic Constraints. Biomimetics 9(1), 59 (Jan 2024). 10.3390/biomimetics9010059

19. Twinanda, A.P., Shehata, S., Mutter, D., Marescaux, J., de Mathelin, M., Padoy, N.: EndoNet: A Deep Architecture for Recognition Tasks on Laparoscopic Videos. IEEE Transactions on Medical Imaging 36(1), 86–97 (Jan 2017). 10.1109/TMI.2016.2593957

20. Ward, T.M., Fer, D.M., Ban, Y., Rosman, G., Meireles, O.R., Hashimoto, D.A.: Challenges in surgical video annotation. Computer Assisted Surgery 26(1), 58–68 (Jan 2021). 10.1080/24699322.2021.1937320

21. Zhu, J., Hamdi, A., Qi, Y., Jin, Y., Wu, J.: Medical SAM 2: Segment medical images as video via Segment Anything Model 2 (Dec 2024). 10.48550/arXiv.2408.00874

